# Costs and outcomes of routine HIV oral pre-exposure prophylaxis implementation across different service delivery models and key populations in South Africa

**DOI:** 10.1101/2023.08.14.23294055

**Authors:** Cheryl Hendrickson, Kamban Hirasen, Constance Mongwenyana, Mariet Benade, Rutendo Bothma, Chantal Smith, Johan Meyer, Brooke Nichols, Lawrence Long

**Affiliations:** Health Economics and Epidemiology Research Office, Faculty of Health Sciences, University of the Witwatersrand, Johannesburg, South Africa; Department of Medical Microbiology, Academic Medical Center, University of Amsterdam, Amsterdam, Netherlands; Department of Global Health, Boston University School of Public Health, Boston, MA, USA; Wits RHI, University of the Witwatersrand, Johannesburg, South Africa; MatCH Maternal, Adolescent and Child Health Institute, KwaZulu-Natal, South Africa; OUT LGBT Well-Being, Johannesburg, South Africa

**Keywords:** South Africa, HIV, pre-exposure prophylaxis, costs and cost analysis

## Abstract

**Background:** Oral pre-exposure prophylaxis (PrEP) is a highly efficacious biomedical HIV prevention tool, yet despite being recommended by the World Health Organization (WHO) since 2015, uptake and persistence remain limited in much of the world, including sub Saharan Africa (SSA). There is a dearth of evidence-based interventions to improve PrEP uptake and persistence in SSA, and the full costs of PrEP programs implemented in routine care settings remain largely unknown. This study aimed to evaluate the cost of delivery of daily oral PrEP, and associated outcomes, to different key and priority populations across different service delivery models (SDMs) in South Africa.

**Methods:** We conducted bottom-up micro-costing of PrEP service delivery from the provider perspective within twelve urban SDMs providing routine PrEP services to various key and propriety populations in Gauteng and KwaZulu-Natal provinces in South Africa. The SDMs included in-facility and outreach models that focused on men who have sex with men (MSM), female sex workers (FSW) and adolescent girls and young women (AGYW). We identified all within- and above-facility activities supporting PrEP delivery, obtained input costs from program budgets, expenditure records and staff interviews, and determined individual resource usage between February 2019 and February 2020 through retrospective medical record review. Our primary outcome was PrEP coverage at six months (defined as having sufficient PrEP drug dispensed at the last visit to be covered at six months post PrEP-initiation). A subset (N=633) of all enrolled subjects had the potential for 12 months of follow-up and were included in a 12-month outcome analysis. We report the cost per client initiated on PrEP in 2021 United States Dollars (USD).

**Findings:** We collected medical record data from 1,281 people who initiated PrEP at 12 SDMs between February and August 2019 and had at least six months of potential follow-up. The average number of visits was 2.3 for in-facility models and 1.5 for outreach models and 3,086 months of PrEP was dispensed. PrEP coverage at six months varied greatly across SDMs, from 41.8% at one MSM-focused fixed clinic to 0% in an MSM-focused outreach model. In general, in-facility programs had higher six-month coverage than outreach programs. Across all SDMs with PrEP clients with potential for 12 months of follow-up (n=633), PrEP coverage at 12 months was 13.6%, with variability between SDMs. The average six-month cost per client initiated on PrEP ranged from $29 to $590, with higher average costs generally observed for the in-facility programs ($152 in-facility versus $84 for outreach). The average monthly cost per PrEP client who had six-month PrEP coverage ranged from $18 to $160 dependent on SDM.

**Interpretation:** This study is an important addition to the PrEP outcome and cost literature in the SSA region. Results show that costs and outcomes vary considerably across different SDMs and populations in real world PrEP programs and provide crucial information for further scale-up of the oral PrEP program in South Africa and the greater SSA region.

## Research in context

### Evidence before this study

Although there have been several modelling studies conducted to investigate the cost and cost outcomes of pre-exposure prophylaxis (PrEP) in sub Saharan Africa, there is a dearth of evidence on the real-world costs of providing PrEP in this region. A search of PubMed conducted before commencement of this study using the terms “PrEP” AND “cost analysis” AND “sub Saharan Africa” for studies published in English between 2016 and 2022 along with the exclusion of publications whose results stemmed from models or that were irrelevant, showed that only nine studies covering four countries have been published that present primary costs of PrEP in this region. Those that have been published generally focus on one population group, are costing particular interventions in a research setting or only use ingredients-based methodology without associated outcomes.

### Added value of this study

We include cost and cost outcomes from a large cohort of PrEP-clients who initiated PrEP across twelve models of care and have at least six months of potential follow-up. Costs are based on actual individual resource utilization data and program expenses, which differentiate it from modelling and ingredients-based costing studies. These results add to the growing body of evidence on the outcomes and associated costs of routine implementation of PrEP programs in the sub Saharan African region, which can be used to strengthen budget impact analyses which will, in turn, assist policy makers in program scale up decisions.

### Implications of all the available evidence

This study provides evidence needed to inform the scale up of PrEP service delivery models. Through the micro-costing approach across several service delivery models, we cost the real-world implementation of these models, which will feed into the evidence needed for HIV prevention scale up and budget planning in South Africa and the wider SSA region. Estimates of provider costs for large scale, routine, effective PrEP delivery platforms that take into account economies of scale, changes in marginal costs as programs expand, understanding the differences in costs and benefits for different populations and risk groups will be vital to the success and sustainability of PrEP programs in SSA.

## Introduction

The HIV epidemic continues to impact millions of people globally, particularly in sub Saharan Africa (SSA), where 60% of new infections occurred in 2020 (1). South Africa is particularly hard hit, with an estimated 210,000 new HIV infections in 2021 and an HIV incidence of 4.19 per 1,000 population (2). Progress on the prevention of HIV infection remains largely stagnant; the global annual number of new infections among adults has hardly changed over the past four years, with the total new infections having declined by 38% from 2.1 million (1.6 million-2.8 million) in 2010 to 1.3 million (1 million-1.7 million) in 2022 (1, 3). This is far short of the 2016 United Nations General Assembly target of 75% for 2020 (4).

Pre-exposure prophylaxis (PrEP) is a highly effective biomedical HIV prevention tool, reducing the risk of HIV-1 acquisition by more than 95% in studies with high adherence (5). However, there are still substantial gaps in the availability of PrEP, with the total number of people using this prevention option at just 28% of the 2020 UNAIDS target of 3 million individuals accessing PrEP in low- and middle-income countries; and just 8% of the new global 2025 target (4, 6). South Africa adopted the World Health Organization’s recommendation to offer PrEP to those at “substantial risk of HIV infection” in 2016, beginning PrEP rollout to select key populations at pilot sites and expanding nationally thereafter (7). Despite this national rollout, PrEP uptake has been slow, with approximately 106,400 people at risk of HIV infection who received PrEP at least once in 2020 (1). This has increased, with almost 350,000 people receiving PrEP in 2021 (2); however, these numbers are still suboptimal. Until recently, the PrEP drug procurement program had been mostly donor funded, however, this is changing such that the South Africa PrEP program is now largely government funded. As South Africa continues to scale-up PrEP provision, data are needed to inform the total cost and affordability of the various service delivery models (SDMs). There is little information, however, to support evidence-based resource allocation decisions in the public sector thus far.

Several modelling studies have investigated the cost-effectiveness of PrEP provision in SSA, and the results are mixed, with large variations in assumed costs of PrEP provision (8–14). Similarly, in South Africa, modelling studies also present results that only show cost-effectiveness under certain conditions such as population-targeted programs, whether PrEP-clients self-select or have high assumed rates of uptake and persistence. These results are largely dependent on the cost estimates used as well as the assumptions around PrEP uptake and persistence (15–20). While modelled estimates provide useful approximations, the most accurate cost estimates are based on actual client resource usage in routine implementation and are critical to determining true cost-effectiveness.

An understanding of the true resource utilization, cost, and expected outcomes of different PrEP service delivery models in South Africa is currently lacking. We have therefore conducted a cost outcomes study to evaluate the cost of delivering daily oral PrEP to different populations across different SDMs in South Africa by estimating the average cost and associated outcomes of PrEP provision in routine settings. These results may serve to inform the expansion of PrEP programs both within South Africa and the greater SSA region and to provide evidence on how to optimize service delivery to improve cost-effectiveness and provide data to conduct budget impact analyses for future policy decisions and implementation strategies in HIV prevention programming.

## Methods

### Study sites and population

We conducted bottom-up micro-costing of PrEP service delivery within twelve urban SDMs providing routine PrEP services in Gauteng and KwaZulu-Natal provinces in South Africa. These models comprised of PrEP services delivered by seven public, multiservice clinics, specialized clinics and community outreach programs, including Technical and Vocational Education and Training colleges (Table 1). Most of the seven sites had both an in-facility and an out-reach model, while some sites were limited to a single model of PrEP provision. The selected SDMs were focused on reaching specific populations considered to be at increased risk for HIV acquisition, including female sex workers (FSW), men who have sex with men (MSM), adolescent girls and young women (AGYW) as well as students at tertiary institutions. All PrEP service delivery was provided per South African national guidelines (21). We intentionally selected SDMs that had been routinely providing PrEP for at least nine months prior to data collection and that had initiated at least 25 people on PrEP with at least six months of potential follow-up prior to data collection.

**Table 1:** Site and service delivery model descriptions.

| Site | Delivery model | Management type | Location | Services offered | Population targeted | PrEP program start date | Total 2019 patient volume | Total 2019 PrEP volume | Proportion PrEP |
| --- | --- | --- | --- | --- | --- | --- | --- | --- | --- |
| **MSM Site 1** | Standalone specialised clinic | NGO | Johannesburg, Gauteng | PrEP, HCT, HIV treatment (ARV) and management, STI screening and treatment, TB screening and referral, Sexual health and lifestyle education and awareness; Community workshops and events; Provision of services at events and venues | MSM | 2019 | 1681 | 716 | 43% |
| **MSM Site 1** | Mobile outreach | NGO | Johannesburg, Gauteng | PrEP, HCT, HIV treatment (ARV) and management, STI screening and treatment, TB screening and referral, Sexual health and lifestyle education and awareness; Community workshops and events; Provision of services at events and venues | MSM | 2019 | 19966 | 728 | 4% |
| **MSM Site 2** | Standalone specialised clinic | NGO | Tshwane, Gauteng | PrEP, HCT, HIV treatment (ARV) and management, STI screening and treatment, TB screening and referral, Sexual health and lifestyle education and awareness; Community workshops and events; Provision of services at events and venues | MSM | 2017 | 4720 | 1908 | 40% |
| **MSM Site 2** | Mobile outreach | NGO | Tshwane, Gauteng | PrEP, HCT, HIV treatment (ARV) and management, STI screening and treatment, TB screening and referral, Sexual health and lifestyle education and awareness; Community workshops and events; Provision of services at events and venues | MSM | 2017 | 3705 | 942 | 25% |
| **FSW Site 1** | Standalone specialised clinic | NGO | Johannesburg, Gauteng | PrEP, ART, STI's, Family Planning, TB and HCT | SW | 2015 | 1339 | 424 | 32% |
| **FSW Site 1** | Mobile outreach | NGO | Johannesburg, Gauteng | PrEP, ART, STI's, Family Planning, TB and HCT | SW | 2015 | 16535 | 1253 | 8% |
| **FSW Site 2** | Standalone specialised clinic | NGO | Tshwane, Gauteng | PrEP, ART, STI's, Family Planning, TB and HCT | SW | 2015 | 632 | 159 | 25% |
| **FSW Site 2** | Mobile outreach | NGO | Tshwane, Gauteng | PrEP, ART, STI's, Family Planning, TB and HCT | SW | 2015 | 10472 | 823 | 8% |
| **FSW Site 3** | Mobile outreach | NGO | Ekurhuleni, Gauteng | PrEP, ART, STI's, Family Planning, TB and HCT | SW | 2015 | 9454 | 858 | 9% |
| **AGYW Site 1** | Community Health Center (CHC) | Government | eThekwini, KwaZulu-Natal | PrEP, ART, SRH, Maternal, Mental health, Youth services and Child Health | AGYW | 2017 | 367811 | 3025 | 1% |
| **AGYW Site 1** | Outreach to TVET campuses | NGO | eThekwini, KwaZulu-Natal | PrEP, ART, SRH, Maternal, Mental health, Youth services and Child Health | Tertiary students | 2017 | 2030 | 2030 | 100% |
| **AGYW Site 2** | Standalone specialised clinic | Government | eThekwini, KwaZulu-Natal | PrEP, ART, SRH, Maternal, Youth services and Child Health | AGYW | 2017 | 47139 | 3294 | 7% |

**Abbreviations:** MSM: Men who have sex with men; SW: Sex worker; AGYW: Adolescent girls and young women; PHC: primary healthcare; TVET: Technical and vocational educational training; NGO: Non-governmental organisation; PrEP: Preexposure prophylaxis; ART: antiretroviral therapy; SRH: Sexual and reproductive health; TB: Tuberculosis; HCT: HIV counselling and testing; STI: Sexually transmitted infection

We enrolled a cohort of clients screened for HIV, found eligible for, and initiated on PrEP, and with at least six months of potential follow-up time between February 2019 to February 2020. We included PrEP-clients who were ≥18 years old and who were recorded as having initiated PrEP within the respective SDM. We excluded PrEP-clients who had missing files, incomplete data, or if they had been transferred out of PrEP care at the selected site within six months of PrEP initiation. Although models focused on specific populations, we did not limit enrolment based on target population. For example, a program focused on MSM may have provided services to a male who did not identify as MSM and this man would have been eligible for inclusion in the costing cohort providing they met the other inclusion/exclusion criteria.

There was no contact with any of the PrEP-clients whose data were used in the study. Data on their medical history, enrolment into PrEP and resource usage during the follow-up period since PrEP initiation was retrospectively extracted from routine client medical records. Within each SDM, we enrolled the most recently initiated PrEP clients who had at least six months of potential follow-up post PrEP initiation. The target enrolment per study population and service delivery model was 200, based on a t-test for sample means, using modelled results of cost estimates for PrEP service delivery in South Africa (16). We estimated that with this sample size, an alpha error rate of 0.05 and 80% power, we would be able to detect at least a 10% difference in cost between SDMs, provided the standard deviation did not exceed 25% of the current modelled cost.

### Data Collection

Routinely collected client-level data from clients’ medical records, including hard copy client files, and both hard copy and electronic client registers, were extracted and managed using REDCap electronic data capture tools hosted at the University of Witwatersrand (22, 23). These data provided individual resource utilization, such as number of clinic visits, service and medications provided, and laboratory tests performed. Facility-level data, such as client headcounts, were extracted from clinic registers. Client-level data for the period February 2019 to February 2020 were collected from September 2020 to March 2021. Facility-level data for the 2019 calendar period were collected from February to August 2021.

### Costing

We estimated the economic costs of PrEP provision from the provider perspective using previously described micro-costing methods (24, 25). We used routine client records to estimate resource utilization for each client over the study period and then multiplied the resource usage by the unit cost. Specific methods and sources for the unit costs for drugs, laboratory tests, clinical staff time, buildings and equipment as well as management and administration costs are shown in Table 2. Resources captured included drugs, diagnostic and laboratory tests, clinical staff time, buildings, equipment, supplies and other shared services, such as utilities and non-clinical staff time. Shared resource costs were collected using facility-level financial records as well as price lists from suppliers and these were proportionally allocated according to the fraction of total visits that were dedicated to PrEP provision. Above site-level management staff salaries were excluded from the main analysis. We collected all costs in South African Rand (ZAR) and inflated the costs to 2021 prices (where necessary) using the South African Consumer Price Index (26). Costs are reported in United States Dollars (USD) using the average 2021 exchange rate of 14.78 ZAR:USD and we assumed a useful life of five years for all capital items (27).

**Table 2:** Costing methods and unit costs.

| Resource | Method for estimating cost | Item | Unit | Cost (USD) | Source |
| --- | --- | --- | --- | --- | --- |
| Drugs | Pre-exposure prophylaxis and other drugs dispensed per PrEP-client during the study period were extracted from the individual client's clinic record. The published national drug unit costs were applied to determine total drug cost per client. | TDF/FTC | Tablet | 0.14 | Master Procurement Catalogue |
| Diagnostic and monitoring laboratory tests | Diagnostic tests included pregnancy tests, urine dipsticks, HIV tests. The actual number of tests performed per PrEP-client during the study period was extracted from the individual client's clinic record. The published unit cost per test was applied to determine the laboratory cost per PrEP-client. | HIV Rapid Screen Test | Test | 3.05 | Department of Health National Laboratory price lists |
|  |  | HIV Serology | Test | 3.82 |  |
|  |  | Alanine Transaminase | Test | 3.14 |  |
|  |  | Hepatitis B Surface Ag Rapid | Test | 8.71 |  |
|  |  | Hepatitis B Surface Ab | Test | 8.71 |  |
|  |  | Urine Dipstick | Test | 0.89 |  |
|  | All laboratory and monitoring tests after PrEP initiation were performed off site by the National Health Laboratory Service. The actual number of tests performed per PrEP-client during the study period was extracted from the individual client's clinic record. The published unit cost per test was applied to determine the laboratory cost per PrEP-client. | RPR* | Test | 1.48 |  |
|  |  | Treponema Hemagglutination | Test | 5.95 |  |
|  |  | Latex Test for Pregnancy | Test | 2.33 |  |
|  |  | BHCG Qualitative | Test | 5.87 |  |
|  |  | Aspartate Transaminase | Test | 3.14 |  |
|  |  | Alanine Transaminase | Test | 3.14 |  |
|  |  | Haemoglobin | Test | 1.25 |  |
| Clinical staff time for PrEP-client care | Clinical staff time was defined as including any doctor, nurse, and HIV lay counselor time spent with the PrEP-client. The clinical staff time cost was calculated by taking the total PrEP clinical staff cost per month and dividing it by the total PrEP visits to the clinic per month to get the average clinical staff cost per visit. Clinical staff salaries were obtained from the site and/or implementing partner salary scales for that cadre. | Not applicable (site specific) | N/A | N/A | Site and/or implementing partner salary scales |
| Buildings and equipment | The total floor space allocated to the PrEP program was calculated by measuring the PrEP care floor space and adding on a proportion of floor space shared by different services (i.e. waiting rooms). A market related average rental cost per square meter was applied to estimate the cost of the building. Electricity costs were obtained for the facility and applied by square meter. Water and effluent costs were not available for the facility and cost per square meter was estimated based on a similar building. The replacement cost of equipment was obtained and a working life of 5 years applied to obtain a cost per month. All PrEP specific equipment was included and a proportion of equipment from shared spaces was included. The building and equipment costs were apportioned to each PrEP-client based on a per-visit cost. | Not applicable (site specific) | N/A | N/A | Site and/or implementing partner financial records |
| Management and administration costs | All staff members who did not provide direct patient care but provided some support to the PrEP program (i.e. clinic manager, data clerks) were included. Staff salaries were obtained from published public sector salaries for that cadre. The cost of all general supplies (i.e. not related to a particular service) was obtained from facility financial reports. Management and administration costs were apportioned to each PrEP-client based on a per visit cost. All costs for management and administration incurred at the site level (i.e. the primary health clinic) were included. Costs incurred above the level of the site were excluded. | Not applicable (site specific) | N/A | N/A | Site and/or implementing partner financial records |

### Outcomes

Our primary outcome (PrEP coverage at six months) for this study was defined as having sufficient PrEP drug dispensed at the last visit to be covered at six months post PrEP-initiation. PrEP outcomes available from routine medical records are difficult to interpret as they do not typically speak directly to adherence. To help understand what is happening in each program we report a number of additional outcomes (same-day initiation, engagement in care, time in care) at various time points, if available. Nine and twelve month outcomes were only reported from the subset of sites that had PrEP clients with 12-months of follow-up. Definitions of each outcome are reported in Table 3.

**Table 3:** Outcome definitions.

| **Outcome** | **Definition** | **Time points** |
| --- | --- | --- |
| Same-day initiation (percentage) | Participant initiated PrEP on the day of presentation for HIV test. Participants that did not initiate on the same day would have had separate visits for HIV testing and PrEP initiation. | Singular |
| Engaged in care after PrEP initiation visit (percentage) | Participant attended at least one visit after PrEP initiation within the follow up period. This is a measure of short-term persistence. Many studies have reported rapid attrition after initiation, which this outcome measures. | Singular |
| Engaged in care (percentage) | Participant attended at least one visit at, or after, specific time point post PrEP initiation. This cannot be reported at 12 months as we did not collect any data related >12 months after PrEP initiation. | 3, 6, 9 months |
| Time in care (days) | Total number of PrEP tablets dispensed during the follow up period. We excluded any tablets dispensed at the last visit which would have covered time outside the follow up period. For example, a participant has 5 visits within 12 months. At the first 4 visits a total of 300 tablets are dispensed. Visit 5 occurs at 330 days and 90 tablets are dispensed, but only 35 of those tablets are counted towards time in care. Time in care for this participant is 335 at 12 months. | 6, 12 months |
| PrEP Coverage | At the PrEP client's last visit in the follow up period, they were dispensed sufficient drug to cover them to the end of the follow up period. | 3, 6, 9, 12 months |

### Data Analysis

All enrolled study participants were included in a six-month cost and outcome analysis. A subset (N=633) of all enrolled subjects across eight SDMs had the potential for 12 months of follow-up and were included in a 12-month outcome analysis. We describe baseline characteristics and outcomes across the study sites, SDM and study population. We also estimated average PrEP-client resource utilization over the six-month study period and the average cost per PrEP-client by SDM and primary outcome and disaggregated by cost category (drugs, laboratory testing, visit costs and fixed costs). Aggregate results are presented in the text, while results disaggregated by site and SDM are reported in the text and tables. Additionally, we also estimated the production cost, a calculation that takes the costs of services to all clients and divides it by only those achieving the primary outcome of PrEP coverage at six months post PrEP-initiation; this incorporates effectiveness into the cost estimates. We also conducted an additional analysis to estimate an average ‘above site’ management cost that can be used as an estimate of costs across the SDMs. All analyses were conducted in Excel (Microsoft Office Standard 2019) using the Healthcare Cost and Outcomes Model to estimate the cost per patient (28) and Rstudio version 1.4.1106.

### Ethics approval

This study was approved by the Human Research Ethics Committee (Medical) of the University of the Witwatersrand (M190621) and the Boston University IRB (H-39120). Data was collected retrospectively and a waiver of informed consent was granted by the ethics review boards.

## Results

### Sample characteristics

We collected data from 1,281 people who initiated PrEP between February and August 2019 across seven sites and 12 SDMs, about half (633/1281) of whom had the potential for 12 months of follow-up across eight of the SDMs (Table 4). This included 1,281 initiation visits and 1,245 follow-up visits amounting to more than 3,000 months of PrEP dispensed. The majority of PrEP-clients were initiated in 2019 (994/1281, 77.6%) while the remainder initiated in 2018. On average, participants were young, with a median age of 27 (IQR: 23-33); this was even lower among the AGYW-focused outreach programs which had a median age of 22 (IQR: 21.0-24.8). About a sixth (63.7%) of PrEP-clients were female with the sex distribution largely influenced by the population the site focused on; however, 8.3% (74/889) of the PrEP initiates at the FSW- and AGYW-focused programs were male.

**Table 4:** Characteristics of PrEP clients at PrEP initiation by site type and service delivery model (n=1,281)

|  |  | MSM Site 1 |  | MSM Site 2 |  | FSW Site 1 |  | FSW Site 2 |  | FSW Site 3 | AGYW Site 1 |  | AGYW Site 2 |
| --- | --- | --- | --- | --- | --- | --- | --- | --- | --- | --- | --- | --- | --- |
|  |  | In-facility (n=98) | Outreach (n=139) | In-facility (n=102) | Outreach (n=53) | In-facility (n=96) | Outreach (n=98) | In-facility (n=42) | Outreach (n=99) | Outreach (n=96) | In-facility (n=168) | Outreach (n=90) | In-facility (n=200) |
| Potential for 12-month follow up | Yes (%) | 97 (99.0) | 71 (51.1) | 0 (0.0) | 0 (0.0) | 93 (96.9) | 98 (100) | 41 (97.6) | 99 (100.0) | 0 (0.0) | 68 (40.5) | 66 (73.3) | 0 (0) |
| Age at PrEP initiation | Mean (SD) | 35.6 (11.5) | 30.4 (8.0) | 32.1 (9.5) | 31.6 (8.3) | 30.3 (6.1) | 29.5 (5.5) | 30.7 (7.6) | 29.7 (6.7) | 28.9 (6.2) | 23.1 (7.5) | 23.3 (5.3) | 25.1 (6.92) |
|  | Median [IQR] | 34 [26, 42] | 29 [25, 35] | 30 [25, 36] | 31 [25, 36] | 30 [26, 35] | 29 [26, 33] | 30 [26, 34] | 29 [25, 34] | 28 [25, 32] | 21 [17, 26] | 22 [21, 25] | 24 [19, 29] |
| Sex | Male (%) | 98 (100.0) | 139 (100.0) | 101 (99.0) | 53 (100.0) | 16 (16.7) | 7 (7.1) | 0 (0.0) | 4 (4.0) | 0 (0.0) | 27 (16.1) | 16 (17.8) | 4 (2.0) |
|  | Female (%) | 0 (0.0) | 0 (0.0) | 1 (1.0) | 0 (0.0) | 80 (83.3) | 91 (92.9) | 42 (100.0) | 95 (96.0) | 96 (100.0) | 141 (83.9) | 74 (82.2) | 196 (98.0) |
| Population<sup>a</sup> | MSM (%) | 92 (93.9) | 61 (43.9) | 94 (92.2) | 10 (18.9) | 6 (6.2) | 4 (4.1) | 0 (0.0) | 4 (4.0) | 0 (0.0) | 0 (0.0) | 0 (0.0) | 1 (0.5) |
|  | FSW (%) | 0 (0.0) | 0 (0.0) | 0 (0.0) | 0 (0.0) | 66 (68.8) | 86 (87.8) | 40 (95.2) | 93 (93.9) | 96 (100) | 0 (0.0) | 0 (0.0) | 2 (1.0) |
|  | AGYW (%) | 0 (0.0) | 0 (0.0) | 0 (0.0) | 0 (0.0) | 2 (2.1) | 0 (0.0) | 0 (0.0) | 2 (2.0) | 0 (0.0) | 111 (66.1) | 51 (56.7) | 110 (55.0) |
|  | PWID, SDC, TGW, TGM or MSW (%) | 2 (2.0) | 0 (0.0) | 3 (2.9) | 1 (1.9) | 0 (0.0) | 3 (3.1) | 0 (0.0) | 0 (0.0) | 0 (0.0) | 0 (0.0) | 1 (1.1) | 0 (0.0) |
|  | Heterosexual males (%) | 1 (1.0) | 40 (28.8) | 2 (2.0) | 0 (0.0) | 2 (2.1) | 0 (0.0) | 0 (0.0) | 0 (0.0) | 0 (0.0) | 19 (11.3) | 5 (5.6) | 3 (1.5) |
|  | Females >24 years of age (%) | 0 (0.0) | 0 (0.0) | 0 (0.0) | 0 (0.0) | 1 (1.0) | 0 (0.0) | 0 (0.0) | 0 (0.0) | 0 (0.0) | 30 (17.9) | 17 (18.9) | 84 (42.0) |
|  | Not indicated (%) | 3 (3.1) | 38 (27.3) | 3 (2.9) | 42 (79.2) | 19 (19.8) | 5 (5.1) | 2 (4.8) | 0 (0.0) | 0 (0.0) | 8 (4.8) | 16 (17.8) | 0 (0.0) |
| Year of PrEP initiation | 2018 (%) | 61 (62.2) | 57 (41.0) | 0 (0.0) | 0 (0.0) | 52 (54.2) | 19 (19.4) | 34 (81.0) | 8 (8.1) | 0 (0.0) | 0 (0.0) | 55 (61.1) | 1 (0.5) |
|  | 2019 (%) | 37 (37.8) | 82 (59.0) | 102 (100.0) | 53 (100.0) | 44 (45.8) | 79 (80.6) | 8 (19.0) | 91 (91.9) | 96 (100.0) | 168 (100.0) | 35 (38.9) | 199 (99.5) |
| Indication for PrEP | Risk assessment by HCW (%) | 34 (34.7) | 6 (4.3) | 81 (79.4) | 9 (17.0) | 42 (43.8) | 65 (66.3) | 9 (21.4) | 33 (33.3) | 77 (80.2) | 120 (71.4) | 52 (57.8) | 165 (82.5) |
|  | Requested by client (%) | 62 (63.3) | 132 (95.0) | 16 (15.7) | 29 (54.7) | 31 (32.3) | 30 (30.6) | 33 (78.6) | 65 (65.7) | 13 (13.5) | 48 (28.6) | 27 (30.0) | 34 (17.0) |
|  | No reason provided (%) | 2 (2.0) | 1 (0.7) | 5 (4.9) | 15 (28.3) | 23 (24.0) | 3 (3.1) | 0 (0.0) | 1 (1.0) | 6 (6.2) | 0 (0.0) | 11 (12.2) | 1 (0.5) |
| Risk assessment completed prior to or at PrEP initiation |  |  |  |  |  |  |  |  |  |  |  |  |  |
|  | Yes (%) | 95 (96.9) | 120 (86.3) | 100 (98.0) | 29 (54.7) | 81 (84.4) | 85 (86.7) | 42 (100.0) | 99 (100.0) | 95 (99.0) | 168 (100.0) | 78 (86.7) | 199 (99.5) |

<sup>a</sup>MSM includes 8 participants who indicated that they are bisexual

In general, the SDMs largely served the population group that they were focusing on (i.e. an MSM-focused program mostly initiated MSM clients on PrEP); however there is variability in population within sites and SDMs. The MSM-focused outreach programs reported that one in five initiates identified as heterosexual men (40/192, 20.8%) compared to the in-facility MSM programs which reported only 1.5% heterosexual males (3/200, 1.5%). Other SDMs showed more heterogeneity in their PrEP-client populations, with the AGYW in-facility sites initiating older women (aged ≥25, 114/368, 31.0%), FSW (2/368, 0.5%), heterosexual males (22/368, 6.0%) and one MSM in addition to the 60.1% AGYW. The AGYW outreach sites were similar, with the majority of PrEP initiates in the AGYW population category (51/90, 56.7%); however, almost one in five persons (17/90, 18.9%) initiated on PrEP were women older than 24 years of age as well as five? (5.6%) heterosexual males. In this AGYW outreach group 16/90 (17.8%) did not indicate which population group they were in. The FSW sites largely initiated FSW (106/138 (76.8%) in-facility and 275/293 (93.9%) outreach); however, a few AGYW, MSM and Transgender women also initiated at these FSW-focused programs.

We observed variability across sites and SDMs on PrEP-clients indication for PrEP, with most sites and SDMs showing a combination of PrEP-clients both initiating PrEP at their request and due to formal risk assessments. However, some were more skewed, for example one MSM-focused outreach model (MSM Site 2) indicated that almost all clients (132/139,95.0%) requested PrEP, while one FSW-focused outreach model (FSW Site 3) had the inverse with the majority (77/96, 80.2%) initiated based on the risk assessment. Within most SDMs, almost all PrEP-clients (1191/1281, 93.0%) had a formal risk assessment done prior to, or at, PrEP initiation. Exceptions to this were the two MSM-focused outreach programs (Site 1: 86.3% and Site 2: 54.7%), the FSW-focused clinic (84.4%) and outreach (86.7%) (Site 3) and the TVET-focused outreach program (Site 6, 86.7%).

### Resource utilization and unit costs

The average six-month resource utilization by site and service delivery model is shown in Table 5. In general, there was higher resource utilization in the in-facility SDMs than in the outreach SDMs. The average number of visits was 2.3 (standard deviation (SD) of 1.7) for the in-facility models and 1.5 (SD: 1.0) for the outreach models. This varied for the specific sites, with MSM Site 2 (in-facility) having the highest average number of visits of 3.0 (SD: 1.5) and the affiliated outreach program having the lowest at 1.0 (SD: 0.1) over the six month period. Across all population groups, the average number of days PrEP was dispensed was higher in the in-facility models (mean: 83, SD: 65) than in the outreach models (mean: 51, SD: 42); this difference between models was greatest among the MSM-focused programs with the outreach programs having the least PrEP dispensed on average (mean: 32; SD: 15) and in-facility having the most (mean: 124; SD: 61). Drug dispensing for sexually transmitted infections and other conditions were also observed, with 121 (9.4%) of PrEP-clients receiving STI treatment and 221 (17.3%) receiving other medication during the first six months post PrEP initiation. Higher proportions of PrEP-clients received STI treatment in the FSW-focused models (14.1-33.3%) compared to the AGYW-focused models (0-4.2%). The mean number of HIV tests administered over the six months post PrEP initiation was 1.87 (SD: 1.22) and 1.43 (SD: 0.84) for in-facility and outreach models respectively. About one creatinine test was done per PrEP-client over the same period, while the other laboratory tests were done less frequently.

**Table 5:** Six-month resource utilisation by site and service delivery model.

|  |  | MSM Site 1 |  | MSM Site 2 |  | FSW Site 1 |  | FSW Site 2 |  | FSW Site 3 | AGYW Site 1 |  | AGYW Site 2 |
| --- | --- | --- | --- | --- | --- | --- | --- | --- | --- | --- | --- | --- | --- |
| 6-month cohort (n=1,281) |  | In-facility<br>(N=98) | Outreach<br>(N=139) | In-facility<br>(N=102) | Outreach<br>(N=53) | In-facility<br>(N=96) | Outreach<br>(N=98) | In-facility<br>(N=42) | Outreach<br>(N=99) | Outreach<br>(N=96) | In-facility<br>(N=168) | Outreach<br>(N=90) | In-facility<br>(N=200) |
| Visits and healthcare worker interactions |  |  |  |  |  |  |  |  |  |  |  |  |  |
| Number of visits | Mean (SD) | 2.7 (1.1) | 1.0 (0.2) | 3.0 (1.5) | 1.0 (0.1) | 1.6 (1.0) | 1.9 (1.0) | 3.1 (2.0) | 2.2 (1.5) | 1.7 (1.0) | 2.1 (1.7) | 1.3 (0.7) | 2.2 (1.9) |
|  | Median [Q1, Q3] | 3 [2, 3] | 1 [1, 1] | 3 [2, 4] | 1 [1, 1] | 1 [1, 2] | 2 [1, 2] | 3 [2, 5] | 2 [1, 3] | 1 [1, 2] | 1 [1, 3] | 1 [1, 1] | 1 [1, 3] |
| Number of nurse interactions | Mean (SD) | 2.7 (1.1) | 1.1 (0.2) | 3.0 (1.5) | 1.1 (0.3) | 1.6 (1.0) | 1.9 (1.0) | 2.9 (1.6) | 2.2 (1.5) | 1.7 (1.) | 2.1 (1.7) | 1.3 (0.7) | 2.2 (1.9) |
|  | Median [Q1, Q3] | 3 [2, 3] | 1 [1, 1] | 3 [2, 4] | 1 [1, 1] | 1 [1, 2] | 2 [1, 2] | 3 [2, 4] | 2 [1, 3] | 1 [1, 2] | 1 [1, 3] | 1 [1, 1] | 1 [1, 3] |
| Number of counsellor interactions | Mean (SD) | 2.6 (1.1) | 1.1 (0.2) | 2.6 (1.2) | 0.9 (0.5) | 1.5 (0.8) | 1.8 (1.0) | 2.7 (1.6) | 2.2 (1.3) | 1.7 (1.0) | 1.3 (1.7) | 1.0 (0.8) | 1.7 (1.6) |
|  | Median [Q1, Q3] | 3 [2, 3] | 1 [1, 1] | 3 [2, 4] | 1 [1, 1] | 1 [1, 2] | 2 [1, 2] | 2 [2, 4] | 2 [1, 3] | 1 [1, 2] | 1 [0, 1] | 1 [0.3, 1] | 1 [1, 2] |
| Drugs, medications or devices dispensed |  |  |  |  |  |  |  |  |  |  |  |  |  |
| Days of PrEP dispensed | Mean (SD) | 130.0 (62.8) | 32.9 (16.9) | 119.0 (59.6) | 30.6 (4.12) | 68.3 (58.9) | 79.6 (63.1) | 79.6 (54.1) | 66.0 (44.3) | 57.8 (42.5) | 65.2 (65.3) | 37.4 (19.7) | 64.5 (55.3) |
|  | Median | 120 | 30 | 120 | 30 | 30 | 59 | 60 | 60 | 30 | 30 | 30 | 30 |
|  | [Q1, Q3] | [120, 186] | [30, 30] | [60, 163] | [30, 30] | [30, 120] | [30, 120] | [30, 120] | [30, 90] | [30, 90] | [30, 90] | [30, 30] | [30, 90] |
| Number of PrEP-clients who had STI medication dispensed<sup>a</sup> | n (%) | 4 (4.1%) | 0 (0.0%) | 15 (14.7%) | 2 (3.8%) | 18 (18.8%) | 19 (19.4%) | 10 (23.8%) | 14 (14.1%) | 32 (33.3%) | 7 (4.2%) | 0 (0.0%) | 0 (0.0%) |
| Number of PrEP-clients who had any other medication dispensed<sup>b</sup> | n (%) | 5 (5.1%) | 0 (0.0%) | 20 (19.6%) | 2 (3.8%) | 26 (27.1%) | 41 (41.8%) | 21 (50.0%) | 42 (42.4%) | 53 (55.2%) | 9 (5.4%) | 2 (2.2%) | 0 (0.0%) |
| Laboratory tests conducted |  |  |  |  |  |  |  |  |  |  |  |  |  |
| HIV test | Mean (SD) | 2.0 (1.0) | 1.0 (0.2) | 2.6 (1.2) | 1.0 (0.2) | 1.4 (0.7) | 1.7 (1.0) | 2.2 (1.2) | 1.9 (1.1) | 1.6 (1.0) | 1.9 (1.5) | 1.3 (0.6) | 1.6 (1.1) |
| Creatinine test | Mean (SD) | 1.2 (1.0) | 1.0 (0.2) | 1.0 (0.7) | 0.2 (0.5) | 1.2 (0.6) | 1.2 (0.6) | 1.6 (0.7) | 1.4 (0.7) | 0.8 (0.5) | 1.0 (0.4) | 0.8 (0.5) | 1.0 (0.3) |
| HBsAg test<sup>c</sup> | Mean (SD) | 0.0 (0.0) | 0.0 (0.0) | 0.2 (0.5) | 0.0 (0.1) | 0.0 (0.1) | 0.0 (0.1) | 0.1 (0.4) | 0.0 (0.2) | 0.0 (0.0) | 0.7 (0.5) | 0.5 (0.5) | 0.0 (0.1) |
| HBsAb test<sup>d</sup> | Mean (SD) | 0.0 (0.0) | 0.0 (0.0) | 0.9 (0.6) | 0.1 (0.3) | 0.8 (0.6) | 0.9 (0.4) | 0.9 (0.5) | 0.8 (0.4) | 0.8 (0.5) | 1.0 (0.3) | 0.7 (0.4) | 1.0 (0.2) |
| Syphilis rapid test | Mean (SD) | 0.0 (0.2) | 0.0 (0.0) | 0.5 (0.9) | 0.1 (0.4) | 0.5 (0.5) | 0.5 (0.5) | 0.9 (0.4) | 0.8 (0.5) | 0.0 (0.1) | 0.9 (0.3) | 0.7 (0.4) | 0.9 (0.3) |
| Pregnancy test | Mean (SD) | 0.0 (0.0) | 0.0 (0.0) | 0.0 (0.0) | 0.0 (0.0) | 0.1 (0.4) | 0.3 (0.8) | 0.5 (0.9) | 0.4 (0.9) | 1.2 (0.9) | 1.3 (1.4) | 0.7 (0.8) | 0.9 (0.4) |
| Other<sup>e</sup> | Mean (SD) | 0.2 (0.4) | 0.0 (0.0) | 0.5 (0.7) | 0.2 (0.4) | 0.2 (0.5) | 0.1 (0.4) | 0.2 (0.4) | 0.1 (0.3) | 0.0 (0.0) | 0.0 (0.2) | 0.1 (0.3) | 0.0 (0.0) |

<sup>a</sup>STI medications include: Ceftriaxone, Cefuroxime, Metronidazole, Doxycycline, Azithromycin, Benzathine Penicillin, Flucloxacillin and Acyclovir.
<sup>b</sup>Other medications include: Paracetamol, Ibuprofen, Diclofenac, Amlodipine, Augmentin, Ciprofloxacin, Bactrim, Allergex, Amoxicillin, Illiadin, Combined Oral contraceptives, Progesterone injection, Saline nasal spray.
<sup>c</sup>Hepatitis B surface antigen
<sup>d</sup>Hepatitis B antibody to surface antigen
<sup>e</sup>Other laboratory tests conducted include: AST, ALT, Hb, Urine dipstix, Papsmear, Hepatitis A IgM, Hepatitis C Antibody, GeneXpert

### PrEP-client outcomes

Overall, the majority (1,191/1,281, 93.0%) of PrEP-clients initiated PrEP the same day as their negative HIV test. One notable exception was the MSM-focused clinic (Site 1), where only a third (32/98, 32.7%) of clients were same-day initiators (Table 6). The proportion of PrEP-clients returning for at least one follow-up visit varied greatly, from only 1.9% at one MSM-focused outreach program to 90.5% at an FSW-focused clinic. Similarly, time in care at six months ranged from an average of 28.9 days (SD: 8.2) to 126 (SD: 61.2) across sites. Our primary outcome, PrEP coverage at six months, varied greatly across SDMs, but was low overall; the highest coverage observed was in an in-facility program focused on reaching MSM (41.8%) while an MSM-focused outreach program had no PrEP-clients with coverage at six months. In general, in-facility programs had higher six-month coverage than the outreach programs, with the exception of one of the FSW-focused programs where the outreach program had a higher coverage (22.4%) compared to the in-facility program (13.5%). The AGYW-focused programs had low coverage, ranging from 4.4% to 15.5% at six months. Three clients (two at an FSW-focused clinic and one at an AGYW-focused clinic) acquired HIV during the study six-month follow-up period.

**Table 6:**
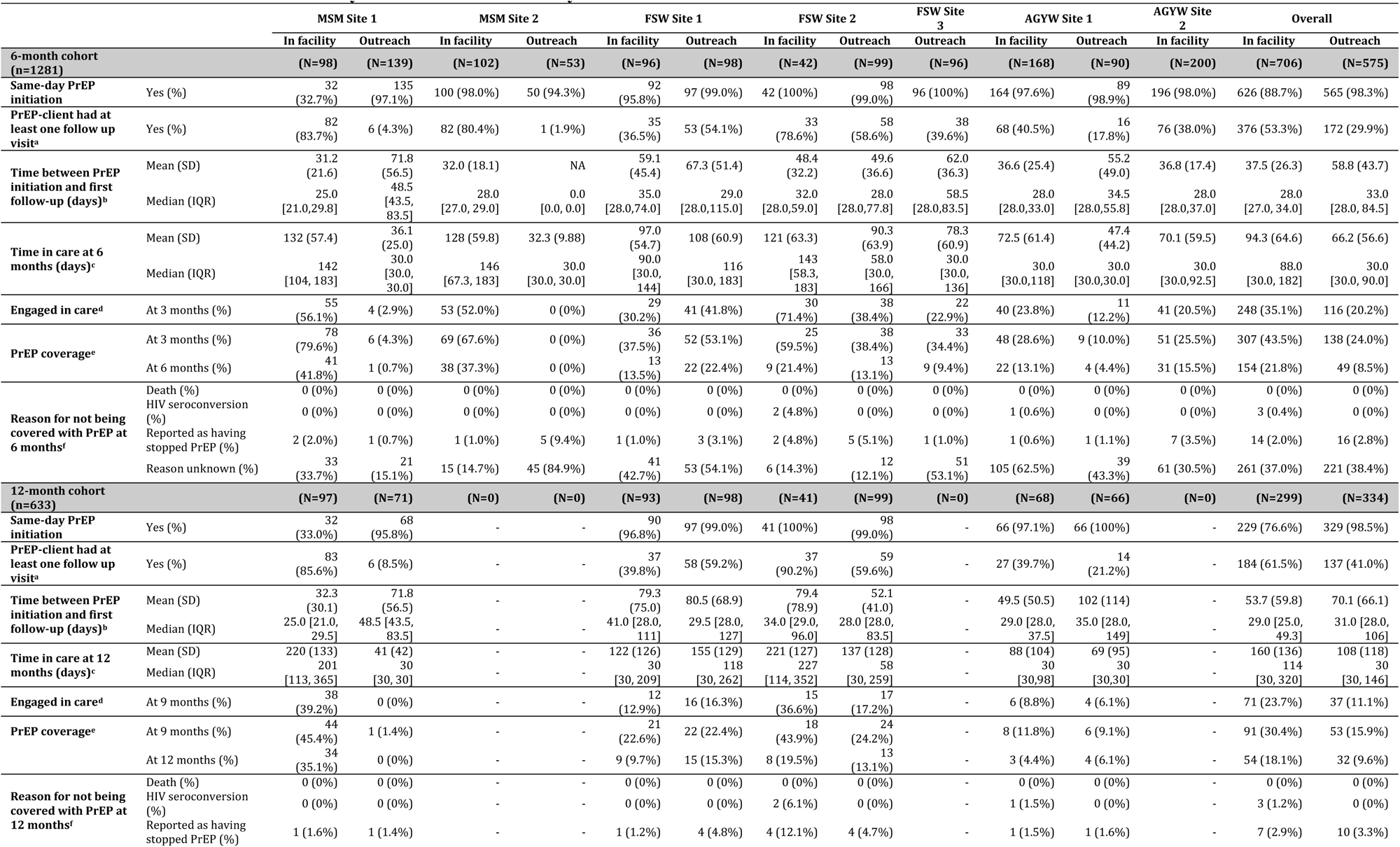
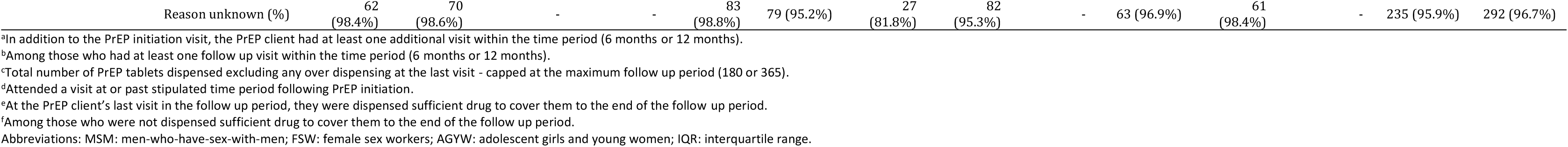
Six- and 12-month outcomes by site and service delivery model.

Similar patterns are observed among the PrEP clients with 12 months of potential follow-up (n=633), with average time in care ranging from 41 days (SD: 42) to 221 (SD: 127). Across all included programs, PrEP coverage at 12 months was 13.6% (86/633), with variability across sites and programs. The MSM-focused clinic had 12-month PrEP coverage of 35.1%, while its accompanying outreach program had no PrEP-clients with this coverage. The two FSW-focused clinics and outreach programs ranged from 9.7% to 19.5% coverage at 12 months, while the AGYW-focused site had 4.4% and 6.1% coverage for the in-facility and outreach programs respectively.

### Costs and cost outcomes

When considering all included clients initiated on PrEP, the average cost per PrEP-client ranged from $29 for an MSM-focused outreach program to $590 for an FSW-focused in-facility program, with higher average costs generally observed for the in-facility programs (Table 7). When estimating the costs for only those who achieved the primary outcome (PrEP coverage at six months), the average cost per PrEP client ranged from $108 at one of the FSW-focused outreach programs to $959 at an FSW-focused in-facility program. Overall, the in-facility programs resulted in higher average costs per PrEP client compared to the accompanying outreach program. This trend held when considering the average cost per PrEP-client among those with PrEP coverage at six months, with the in-facility programs ranging from $159 to $920 and the outreach programs from $108 to $220 over six months. While the average costs might have been lower for outreach programs, the production costs, or six-month costs to produce one client with six-month PrEP coverage were, on average, much higher. The exception to this were the FSW-outreach programs which had lower production costs than the corresponding in-facility programs.

**Table 7:** Estimated costs per PrEP-client of six months of PrEP provision by site and service delivery model. Costs reported in 2021 USD (95% CI).

|  | MSM Site 1 |  | MSM Site 2 |  | FSW Site 1 |  | FSW Site 2 |  | FSW Site 3 | AGYW Site 1 |  | AGYW Site 2 |
| --- | --- | --- | --- | --- | --- | --- | --- | --- | --- | --- | --- | --- |
| Service delivery model | In-facility | Outreach | In-facility | Outreach | In-facility | Outreach | In-facility | Outreach | Outreach | In-facility | Outreach | In-facility |
| **Proportion of clients with PrEP coverage at 6 months** | 41/98<br>(41.8%) | 1/139<br>(0.7%) | 38/102<br>(37.3%) | 0/53<br>(0%) | 13/96<br>(13.5%) | 22/98<br>(22.4%) | 9/42<br>(21.4%) | 13/99<br>(13.1%) | 9/96<br>(9.4%) | 22/168<br>(13.1%) | 4/90<br>(4.4%) | 31/200<br>(15.5%) |
| **Average 6-month cost per PrEP-client** |  |  |  |  |  |  |  |  |  |  |  |  |
| All PrEP-clients | 228<br>(208-248) | 76<br>(73-79) | 191<br>(175-208) | 29<br>(26-31) | 125<br>(111-140) | 75<br>(68-83) | 590<br>(488-692) | 105<br>(93-117) | 131<br>(116-147) | 84<br>(76-92) | 61<br>(55-68) | 73<br>(66-80) |
| PrEP clients with PrEP coverage at 6 months | 312<br>(293-332) | 140 | 248<br>(236-262) | - | 236<br>(204-268) | 108<br>(92-125) | 959<br>(753-1164) | 220<br>(190-250) | 276<br>(231-320) | 179<br>(155-203) | 132<br>(43-220) | 170<br>(160-181) |
| **Average cost per person-month per PrEP-client<sup>a</sup>** |  |  |  |  |  |  |  |  |  |  |  |  |
| All PrEP-clients | 58<br>(53-63) | 71<br>(70-73) | 53<br>(49-58) | 28<br>(26-31) | 68<br>(62-74) | 32<br>(29-34) | 182<br>(159-204) | 44<br>(41-47) | 66<br>(62-70) | 47<br>(42-52) | 48<br>(45-51) | 37<br>(36-39) |
| PrEP clients with PrEP coverage at 6 months | 52<br>(49-55) | 23 | 142<br>(39-44) | - | 39<br>(34-45) | 18<br>(15-21) | 160<br>(126-195) | 37<br>(32-42) | 46<br>(39-53) | 30<br>(26-34) | 22<br>(7-39) | 28<br>(27-30) |
| **Cost breakdown for PrEP-clients with PrEP coverage at 6 months** |  |  |  |  |  |  |  |  |  |  |  |  |
| Drugs | 26<br>(24-28) | 5 | 24<br>(23-26) | - | 13<br>(13-13) | 18<br>(14-22) | 20<br>(13-26) | 21<br>(17-25) | 16<br>(11-21) | 24<br>(17-31) | 10<br>(1-19) | 24<br>(22-26) |
| Lab tests | 16<br>(14-19) | 12 | 35<br>(30-39) | - | 31<br>(24-38) | 26<br>(21-30) | 32<br>(28-37) | 33<br>(27-39) | 28<br>(23-34) | 43<br>(38-49) | 35<br>(6-63) | 28<br>(25-30) |
| Clinic staff visit cost | 133<br>(124-142) | 47 | 127<br>(120-135) | - | 89<br>(77-102) | 15<br>(13-17) | 744<br>(573-915) | 60<br>(51-69) | 23<br>(19-28) | 69<br>(56-82) | 45<br>(17-73) | 61<br>(57-65) |
| Fixed costs (buildings, equipment, management, admin) | 136<br>(128-146) | 75 | 63<br>(58-67) | - | 103<br>(89-117) | 50<br>(43-57) | 163<br>(130-196) | 107<br>(90-124) | 208<br>(170-245) | 42<br>(37-48) | 42<br>(16-67) | 58<br>(54-62) |
| **Production cost of PrEP coverage at 6 months** | 545 | 10,537 | 514 | - | 929 | 336 | 2,754 | 780 | 1,403 | 643 | 1,379 | 470 |

<sup>a</sup> Total cost per month divided by the total number of PrEP tablets dispensed excluding any over dispensing at the last visit - capped at the maximum follow up period (180 or 365).
<sup>b</sup> Above-site costs were collected for two sites and an 'above-site cost' was estimated for each PrEP-client in our sample at these sites. We then averaged this amount to determine an average 'above-site cost' for each PrEP-client overall and applied this cost over-and-above the costs collected at the other sites.

## Discussion

In this micro-costing study, we estimated the real-world outcomes and costs of PrEP provision across seven sites that made up 12 SDMs reaching MSM, FSW and AGYW populations. The costs of six months of PrEP provision varied widely across sites and service delivery models from $29 to $590 per client initiated and $108 to $959 among those who achieved PrEP coverage at six months. These costs were largely driven by clinic personnel and fixed costs, which is similar to costs published from other studies in the region (29–31). Meyer-Rath et al. (32) used an ingredients-based approach relying on expert opinion, available literature and several assumptions to estimate the cost per client year for the first year on PrEP for young women in South Africa, obtaining an estimate of approximately US$124; no specific SDM was indicated. This is generally lower than estimated through this costing of real-world service delivery, highlighting the importance of micro-costing in providing a more granular view of costs and associated outcomes of PrEP provision, which is key for future program, intervention and budget planning. When compared to the published costs of providing PrEP in other micro-costing studies done in the region, the costs observed in these SDMs are generally higher, with annual costs ranging from $92 to $943 per year in these studies (31, 33–39). Two possible explanations for these differing results are the more extensive inclusion of programmatic costs and relatively low PrEP client volumes in the SDMs with particularly high costs. Some of the SDMs were still within their first year or two of PrEP provision and this may have contributed to both low client volumes and low PrEP persistence as models were still learning how to effectively promote PrEP among those who might benefit from it.

Overall, in-facility programs were costlier per person on PrEP than the outreach programs, however, they generally had better PrEP coverage at six months, resulting in lower production costs. A notable exception to this were the FSW outreach programs which had lower production costs than the in-facility programs, highlighting the need for differentiated programs that focus on PrEP-client needs – in this case, PrEP service delivery offered at the FSWs places of work. Some of the high costs can be attributed to the low PrEP-client volume and we observe patterns of lower costs with higher client volumes. This could indicate potential for economies of scale for these programs.

Across all sites and service delivery models we observed low PrEP coverage over time, ranging from 0% to 50% at six months post PrEP initiation. This is in line with other literature from the region (31, 40). This is concerning given that these groups are disproportionally affected by HIV. AGYW accounted for 25% of new HIV infections in 2020, despite representing just 10% of the population in SSA with an estimated 4,200 AGYW infected with HIV every week in 2020 (1, 6). Additionally, in 2020, key populations (sex workers and their clients, gay men and other men who have sex with men, people who inject drugs, transgender people) and their sexual partners accounted for 65% of HIV infections globally, and accounted for 39% of new HIV infections in sub-Saharan Africa (6). The associated costs presented here will be helpful in future budget planning for interventions aimed at improving the low PrEP coverage among populations at risk of HIV acquisition in the SSA region.

There are several limitations in this study. Firstly, the limited information available around ongoing risk in the PrEP cohort limits any conclusions about who is persisting versus discontinuing on PrEP. Risk assessments were not captured at all visits and the reasons for stopping PrEP and changes in risk cannot be determined. Therefore, apart from three clients who seroconverted during the six month follow-up period, it is not known whether PrEP-client changes in outcomes were due to changes in risk, adoption of other HIV prevention mechanisms or simply discontinuation of PrEP with no change in HIV acquisition risk. Additionally, the documentation of risk or reason for PrEP initiation at first visit was not standardized and, as such we are unable to say whether these programs were initiating the people who would most benefit from being on PrEP. We were also not able to trace the movement of clients from one site or model to another. We therefore are not able to accurately distinguish transfer of PrEP care to another facility from loss-to-follow-up. With regards to costing, demand creation efforts were not captured in a systematic and measurable way so we are unable to differentiate those costs from the broader fixed costs of the program. Additionally, we only captured resource utilization from the point of PrEP initiation, so we are unable to attribute any costs associated with PrEP demand creation and screening at the individual level. It is becoming increasingly clear that these are important components of PrEP provision programs that need to be included when considering PrEP program scale-up.

Despite these limitations, our study is an important addition to the PrEP cost literature in the SSA region, providing vital outcome and unit cost data on real world PrEP provision in South Africa across a diverse set of service delivery models and populations. These data may be used to strengthen budget impact and cost-effectiveness analyses, providing crucial information for further scale-up of the oral PrEP program in South Africa and the greater SSA region.

## Data Availability

All data produced in the present study are available upon reasonable request to the authors.

## Acknowledgements

We are grateful to the men and women who contributed their data to this study. We thank the implementing partners and clinic staff for their time and efforts in providing us with the data required to carry out this analysis. Additionally, we thank the study staff (Nkamoheleng Mokhesi, Nonhlanhla Tshabalala, Portia Ngwenya, William Magolego and Lerato Molapo) for their excellent assistance in data collection.

## Funding

This study has been made possible by the generous support of the American People and the President’s Emergency Plan for AIDS Relief (PEPFAR) through US Agency for International Development (USAID) under the terms of Cooperative Agreement 72067419CA00004 to HE^2^RO. CH was supported by the Fogarty International Center and National Institute of Mental Health through the National Institutes of Health award number D43 TW010543. LL was supported by the National Institute of Mental Health of the National Institutes of Health award number K01MH119923. The contents are the responsibility of the authors and do not necessarily reflect the views of PEPFAR, USAID, NIH or the US Government.

## Competing interests

All authors declare that they have no competing interests.

## Author contributions

CH and LCL led conception of the manuscript, design as well as drafting the manuscript, data analysis and interpretation of results with contributions from KH, CM and MB. KH, CM and MB contributed to the acquisition of the data, analysis and interpretation of the data and critical revision of the manuscript. RB, JM, and CS contributed by providing site and resource utilization data. BN contributed in the critical revision of the manuscript. All authors reviewed and approved the final version of the manuscript.

## Data sharing statement

The underlying individual level patient resource data and costs remain the property of the study sites. On application to the study team specific site level and ethics approval can be obtained as required.

## Figure legends

No figures included

## Notes

**Supported by:** USAID 72067419CA00004 LL time NIMH K01MH119923 CH time D43 TW010543

### Competing Interest Statement

The authors have declared no competing interest.

